# Dental Anxiety in Oral–Systemic Health: Empirical Evaluation of a Published Conceptual Model

**DOI:** 10.64898/2026.09.25.26364045

**Authors:** Mika Kajita, Akie Yada, Pekka Ylöstalo, Vesa Pohjola, Kirsi Sipilä, Gerald Humphris, Juha Auvinen, Satu Lahti

## Abstract

**Objectives:** A previously published conceptual mapping review integrated fragmented evidence on dental anxiety (DA) into a broader conceptual model of potential causes and consequences. This study focused on consequences, extending from oral health-related behaviours through oral disease to systemic health, and empirically evaluated these hypothesized relationships on the population level.

**Methods:** Cross-sectional data from the 46-year follow-up of the Northern Finland Birth Cohort 1966 (n = 1,859) were analysed using structural equation modelling. DA was modelled as a latent construct; delayed/problem-oriented dental attendance and home oral care as behavioural proxies; periodontal disease burden as the percentage of sites with probing pocket depth ≥4 mm; and cardiovascular risk as a Framingham-based risk score. Generalized anxiety, sex, and education were covariates. Smoking-stratified multigroup analyses assessed model robustness.

**Results:** The prespecified associations were observed across behavioural, periodontal, and systemic-risk domains. Higher DA was associated with more delayed/problem-oriented attendance and poorer home care; both were associated with greater periodontal disease burden, which in turn was associated with higher cardiovascular risk. Model fit was good (χ²(119) = 529.98, CFI = 0.962, TLI = 0.951, RMSEA = 0.043, SRMR = 0.051). The overall association between DA and cardiovascular risk was small (β = 0.066, p = .004), with a small decomposed indirect association (β = 0.013, p = .006) and a remaining direct association (β = 0.053, p = .022). Smoking-stratified multigroup comparisons found no differences in the structural associations by smoking status.

**Conclusions:** This population-based evaluation provides preliminary empirical support for the conceptual model. Findings extend the relevance of DA beyond dental attendance and oral health outcomes to a broader oral–systemic public-health context. Cross-sectional data cannot establish temporal or causal ordering; longitudinal replication is needed.

## Introduction

Dental anxiety (DA) is a common oral health-specific psychosocial factor, affecting around 15% of adults [1]. It has been consistently associated with poorer oral health-related behaviours, especially disrupted dental attendance, and adverse oral health outcomes [2, 3, 4]. Longitudinal studies have shown that increased DA leads to avoidance of oral health care [5, 6]. These associations place DA within a broader public-health context in which psychosocial and behavioural factors are relevant to oral healthcare use and oral health [7, 8].

Oral health-related behaviours are associated with periodontal health, and periodontal disease has established associations with systemic diseases, particularly cardiovascular disease [9, 10]. A previous conceptual mapping review integrated existing models of the determinants and consequences of DA and identified a specific gap: systemic health had rarely been incorporated into DA-focused frameworks [4]. The review therefore proposed a broader conceptual model spanning psychological and environmental determinants, DA, oral health-related behaviours, oral disease, and systemic health, and identified empirical evaluation as the next step.

Empirical evaluation of a conceptual model requires its constructs and hypothesized relationships to be operationalized using measurable variables. Baker et al. emphasized the need to evaluate and adapt conceptual models in oral health research and specific settings [11]. Celeste et al. further discussed the challenge of operationalizing conceptual models using measurable variables, particularly when existing datasets capture only part of a broader conceptual framework [12]. The age-46 NFBC1966 survey wave provided a particularly suitable setting for this operationalization because DA and oral health-related behaviours were assessed alongside clinical oral and medical examinations within the same population-based cohort.

Accordingly, the aim of this study was to provide the first empirical evaluation of the consequences component of the previously proposed conceptual model by operationalizing its key behavioural, periodontal, and systemic disease-risk domains in the NFBC1966 and testing the hypothesized associations using cross-sectional structural equation modelling (SEM; Figure 1). The prespecified hypotheses were that higher DA would be associated with more delayed/problem-oriented oral healthcare attendance and poorer home oral care; that these behavioural patterns would be associated with greater periodontal disease burden; and that greater periodontal disease burden would be associated with higher cardiovascular risk. A further hypothesis was that DA would show an overall positive association with cardiovascular risk, including an indirect component across the measured behavioural and periodontal domains.

**Figure 1.**
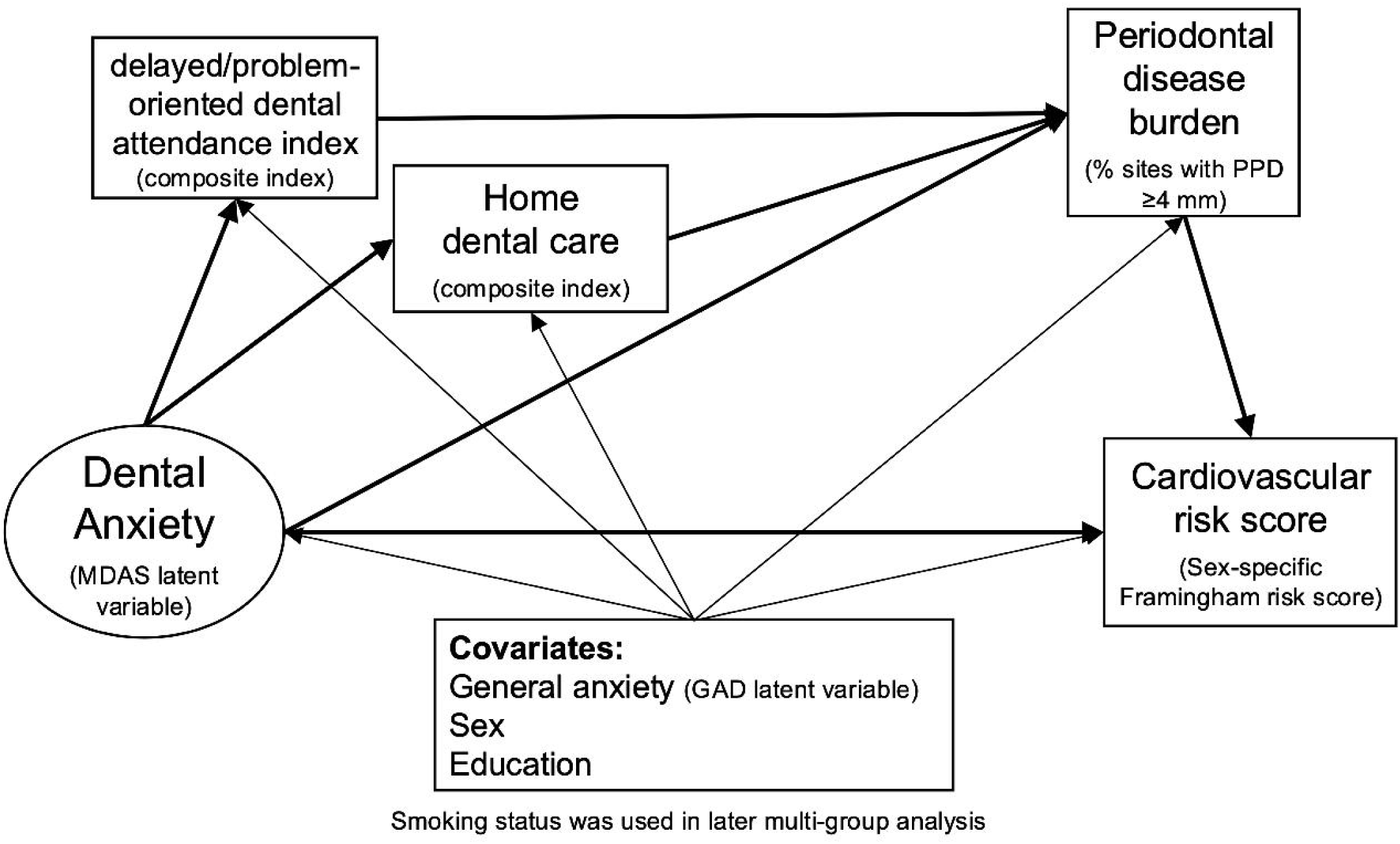
Hypothesized operational model used to evaluate the previously proposed conceptual model in the present study.

## Methods

### Study design and sample

The Northern Finland Birth Cohort 1966 (NFBC1966) is a population-based birth cohort that originally included children with expected dates of birth in 1966 in the two northernmost provinces of Finland (n = 12,231) [13]. The present study was a cross-sectional analysis of the 46-year follow-up conducted in 2012–2013. Cohort members living within 100 km of Oulu were invited to clinical oral and medical examinations (n = 3,150); 1,964 (62.3%) attended the dental examination, and 1,923 with valid periodontal examination data comprised the analytic sample. Participants in the 46-year follow-up were more often female, employed, married, and of higher education and social class than non-participants [14]. All participants provided written informed consent. The study was conducted in accordance with the Declaration of Helsinki and approved by the Ethical Committee of the Northern Ostrobothnia Hospital District (74/2011). As this was a secondary analysis, no formal a priori sample size calculation was performed.

### Conceptual model and operationalization

The operational model shown in Figure 1 was derived from the previously published conceptual model [4]. The behavioural, periodontal, and systemic-risk domains were operationalized using variables available at the 46-year NFBC1966 follow-up. The hypothesized ordering was specified a priori based on the published model and prior evidence; because the analysis was cross-sectional, the directional arrows indicate hypothesized ordering rather than established temporal or causal relationships.

### Measures

DA was measured using the Modified Dental Anxiety Scale (MDAS), a valid and reliable five-item instrument for self-rating DA translated into Finnish [15, 16]. Items are rated from 1 (not anxious) to 5 (extremely anxious), yielding a total score of 5–25. In the present sample, McDonald’s ω was 0.918.

### Oral health behavior

As the NFBC1966 dataset did not include established measures of the relevant behavioral domains, two a priori composite indicators were constructed as pragmatic theory-based proxies: delayed/problem-oriented dental attendance and home oral care. These composites were not intended as reflective psychometric scales; rather, they combined distinct behavioral aspects selected a priori to operationalize the corresponding conceptual domains. The delayed/problem-oriented dental attendance index combined time since the last dental visit, as an indicator of delayed or infrequent attendance, with pain at the last dental visit, to capture problem-oriented attendance. These components were selected to represent these distinct aspects of dental attendance rather than on the basis of their intercorrelation. Painful dental experiences may reinforce DA and contribute to delayed re-attendance [2, 17]. The home oral care index combined floss use, interdental brush use, toothbrushing frequency, and skipping toothbrushing because of fatigue or lack of motivation. The attendance index was calculated as the mean of the available recoded values for time since the last visit and pain at the last visit, whereas the home oral care index was calculated as the mean of four binary items when at least three were available. Higher scores indicated more delayed/problem-oriented attendance or better home oral care, respectively. Detailed coding is provided in Appendix Table 1.

### Periodontal disease

Periodontal examinations were conducted by seven calibrated dentists using a mouth mirror and a ball-pointed periodontal probe with 2-mm gradation (LM 8-520B, Lääkintämuovi, Finland). Probing pocket depth (PPD) was measured at four sites per tooth (mesiobuccal, midbuccal, distobuccal, and midoral) in all teeth except third molars and residual roots. The probing force (25 g) was calibrated before each examination [18]. For the present study, periodontal disease burden was operationalized as the percentage of examined sites with PPD ≥4 mm per participant.

### Cardiovascular risk

Cardiovascular risk was assessed using a previously derived sex-specific 2008 Framingham risk score variable available in the NFBC1966 dataset [19], based on the equation of D’Agostino et al [20]. The algorithm incorporates age, smoking status, systolic blood pressure, antihypertensive treatment, total cholesterol, high-density lipoprotein (HDL) cholesterol, and diabetes status. In the present study, this score was used as a continuous observed outcome variable.

### General anxiety

General anxiety symptoms were assessed using the 7-item Generalized Anxiety Disorder scale (GAD-7) [21]. The measure asks how often respondents were bothered by 7 anxiety-related symptoms during the previous 2 weeks, with response options ranging from 1 (“not at all”) to 4 (“nearly every day”) in the source dataset. These item-level responses were used directly as indicators of a latent general anxiety construct in the SEM. For descriptive reporting and comparability with previous studies, responses were recoded to the conventional 0–3 scale and summed to yield a total score ranging from 0 to 21. In the present sample, McDonald’s ω for the GAD-7 was 0.893.

### Covariates and stratification

Education, sex, and general anxiety were included as covariates because they have been associated with DA [22, 23, 24] and with coronary heart disease or cardiovascular risk [20, 25, 26]. Smoking was considered separately because it is relevant across several model domains [27, 28] and is a component of the Framingham risk score; it was therefore not included as an additional covariate. Robustness with respect to smoking was examined using smoking-stratified sensitivity analyses.

Sex was obtained from birth records. Education was classified as (1) less than 9 years of basic education, (2) completed basic education, or (3) upper secondary education. Smoking was dichotomized as current or non-current. Current smokers included those who smoked occasionally or at least weekly; non-current smokers included never smokers, those with a smoking history of less than 1 year, and former smokers who had quit at least 1 year earlier after smoking at least one cigarette per day for more than 1 year.

### Statistical Analyses

Descriptive statistics and missingness patterns were examined. Little’s MCAR test [29] assessed whether the data were consistent with missing completely at random. Missing data in endogenous variables and latent indicators were handled using full information maximum likelihood under a missing-at-random assumption; participants with missing education were excluded from the covariate-adjusted SEM.

The MDAS and GAD-7 were evaluated using confirmatory factor analysis. SEM then tested the hypothesized model shown in Figure 1. DA and general anxiety were modelled as latent variables; the behavioural, periodontal, and cardiovascular variables were observed, with education and sex included as observed covariates. The model estimated total, remaining direct, and model-based indirect associations between DA and cardiovascular risk. Because the data were cross-sectional, indirect associations were interpreted as decomposed associations rather than evidence of causal mediation.

Because the MDAS items and cardiovascular risk score were non-normally distributed, robust maximum likelihood estimation was used. Model fit was assessed using the chi-square statistic, CFI, TLI, RMSEA, and SRMR, with conventional thresholds for acceptable fit (CFI/TLI ≥0.90; RMSEA ≤0.08) and good fit (CFI/TLI ≥0.95; RMSEA ≤0.06; SRMR ≤0.08) [30, 31].

For smoking-stratified analyses, measurement invariance of the MDAS and GAD-7 was assessed at the configural, metric, and scalar levels. Practical invariance was evaluated primarily using ΔCFI ≤0.010 [32], with scaled chi-square difference tests also reported. Between-group differences in structural paths, indirect associations, and the total association were then tested.

Analyses were conducted using Mplus version 8.11 (Muthén & Muthén, Los Angeles, CA, USA); all tests were two-sided. Reporting followed the Strengthening the Reporting of Observational Studies in Epidemiology (STROBE) guidelines.

### Use of generative artificial intelligence

Generative artificial intelligence (ChatGPT, OpenAI) was used to assist with literature searching, brainstorming, manuscript drafting and language editing, drafting and checking Mplus syntax, and preparation of Zenodo repository materials. Literature identified with AI assistance was verified against the original sources, and all AI-assisted text, code, and repository materials were reviewed and revised by the authors. AI was not used to perform statistical analyses or make analytical or interpretative decisions.

## Results

Table 1 summarizes the characteristics of the study sample (n = 1,923). Item-level descriptive statistics for the MDAS and GAD-7 and the distributions of the behavioral indicators are provided in Appendix Tables 2 and 3. Time since the last dental visit and pain at the last dental visit were weakly correlated, indicating limited overlap between the two components (Appendix Table 4). Missingness was generally limited but somewhat higher for the GAD-7 items and cardiovascular risk score (Appendix Table 5). Little’s missing completely at random (MCAR) test was χ²(298) = 441.65, p < .001, indicating that the strict MCAR assumption was not supported.

**Table 1.** Sample characteristics and descriptive statistics of the study variables.

| Variable | n (%) or mean ± SD | Missing, n (%) |
| --- | --- | --- |
| Sex |  | 0 |
| Male | 892 (46.4) |  |
| Female | 1031 (53.6) |  |
| Education |  | 64 (3.3) |
| Less than 9 years of basic education | 7 (0.4) |  |
| Completed basic education | 979 (50.9) |  |
| Upper secondary education | 873 (45.4) |  |
| Smoking status |  | 62 (3.2) |
| Non-current smoker | 1,442 (75.0) |  |
| Current smoker | 419 (21.8) |  |
| Dental anxiety |  | 51 (2.7) |
| MDAS total score | 9.26 ± 3.99 | |
| Anxiety symptoms |  | 251 (13.1) |
| GAD-7 total score | 2.42 ± 3.11 | |
| Oral health behavior |  |  |
| Delayed/problem-oriented dental attendance index | 0.74 ± 0.27 | 51 (2.7) |
| Home care index | 0.56 ± 0.26 | 132 (6.9) |
| Periodontal disease burden |  | 26 (1.4) |
| Percentage of sites with probing pocket depth ≥4 mm | 2.76 ± 6.26 | |
| Cardiovascular risk |  | 179 (9.3) |
| Framingham risk score | 6.37 ± 4.79 | |

MDAS = Modified Dental Anxiety Scale; GAD-7 = 7-item Generalized Anxiety Disorder scale. The delayed/problem-oriented dental attendance index and home care index were composite indicators derived from multiple observed variables; details of their construction are provided in the Appendix Table 1. Higher values indicate a greater tendency toward delayed/problem-oriented dental attendance and better home care, respectively. Periodontal disease burden was defined as the percentage of sites with probing pocket depth ≥ 4 mm. Cardiovascular risk was assessed using the Framingham risk score.

The initial one-factor CFA of the MDAS showed suboptimal fit. Allowing a correlated residual between MDAS1 and MDAS2, which both assess anticipatory DA, substantially improved fit; this specification was retained in subsequent analyses (Appendix Table 6) [33]. The one-factor CFA of the GAD-7 showed acceptable fit, with moderate to high standardized factor loadings (Appendix Table 7).

After excluding 64 participants with missing education data, the covariate-adjusted structural model included 1,859 participants and showed good fit according to the practical fit indices, χ²(119) = 529.98, p < .001, CFI = 0.962, TLI = 0.951, RMSEA = 0.043, and SRMR = 0.051. As shown in Figure 2, the hypothesized pattern of associations was observed across the behavioral, periodontal, and systemic-risk domains. Higher DA was associated with more delayed/problem-oriented dental attendance and poorer home oral care. These behavioral indicators were associated with greater periodontal disease burden, which was associated with higher cardiovascular risk score. The total association of DA with cardiovascular risk score was β = 0.066, p = .004. The decomposed indirect association was small β = 0.013, p = .006, and a remaining direct association was observed (β = 0.053, p = .022) (Table 2).

**Figure 2.**
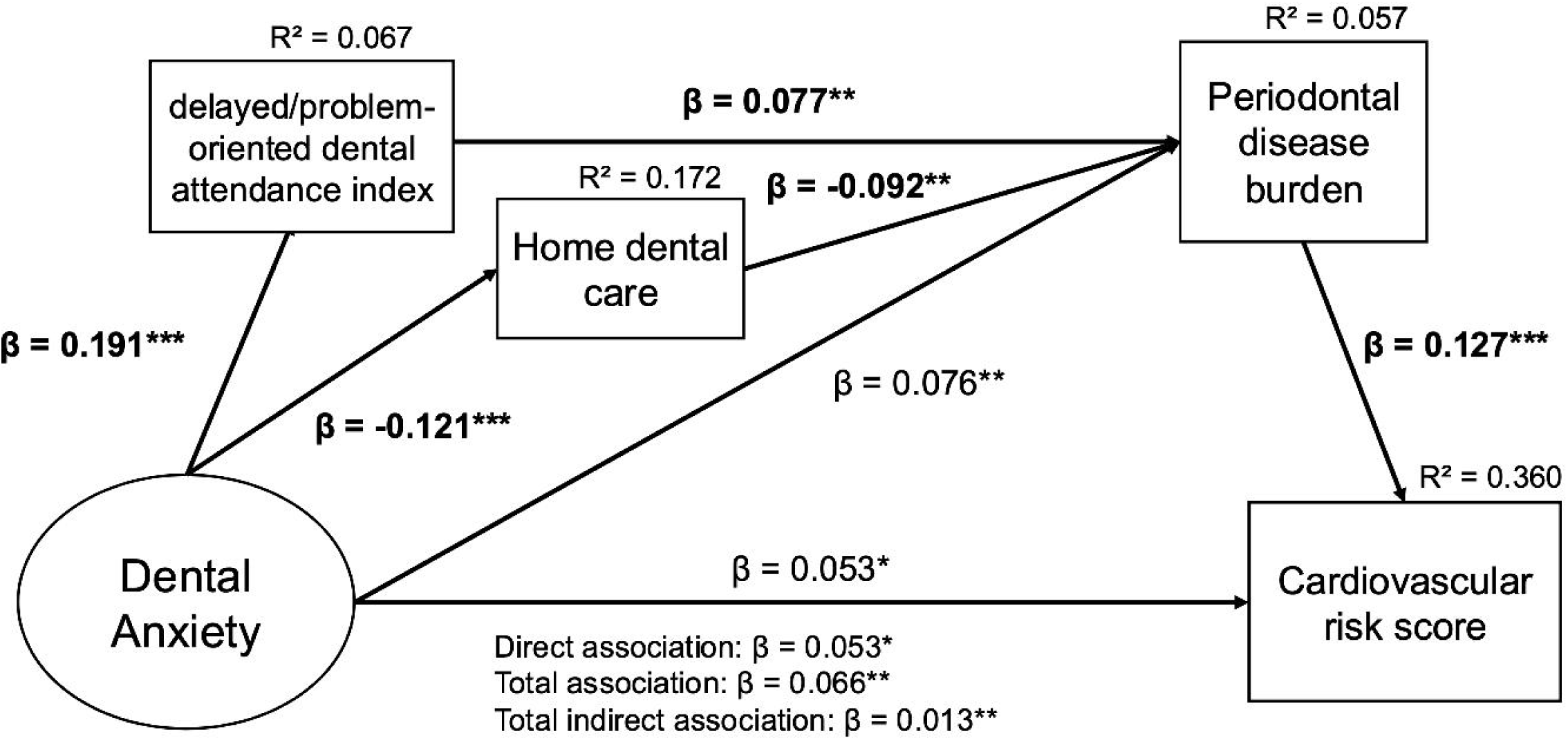
Estimated prespecified structural equation model of dental anxiety, oral health-related behaviours, periodontal disease burden, and cardiovascular risk. *Arrow labels indicate standardized estimates (STDYX), and values above endogenous variables indicate explained variance (R²). The model included a residual covariance between MDAS1 and MDAS2 and was adjusted for generalized anxiety, sex, and education; covariate paths are not shown. Model fit was* χ*²(119) = 529.98, CFI = 0.962, TLI = 0.951, RMSEA = 0.043 (90% CI, 0.039 to 0.047), and SRMR = 0.051. Direct, indirect, and total associations between dental anxiety and cardiovascular risk are reported in Table 2. *p < .05, **p < .01, ***p < .001*.

**Table 2.** Total, direct, and indirect associations of dental anxiety with cardiovascular risk score.

| Association | Standardized $\beta$ | 95% CI | p-value |
| --- | --- | --- | --- |
| Total association | 0.066 | 0.021 to 0.111 | .004 |
| Direct association | 0.053 | 0.008 to 0.098 | .022 |
| Total indirect association | 0.013 | 0.004 to 0.022 | .006 |
| Indirect via periodontal disease burden alone | 0.010 | 0.001 to 0.018 | .023 |
| Indirect via delayed/problem-oriented and periodontal disease burden | 0.002 | 0.000 to 0.003 | .022 |
| Indirect via home dental care and periodontal disease burden | 0.001 | 0.000 to 0.003 | .027 |

**Note:** Values are standardized estimates (STDYX) from a structural equation model using robust maximum likelihood (MLR). The model included a residual covariance between MDAS1 and MDAS2 and was adjusted for generalized anxiety, sex, and education. Indirect associations were estimated as model-based products of coefficients in the prespecified SEM. Because the data are cross-sectional, these estimates should be interpreted as decomposition components of association, not as evidence of causal mediation or temporal pathways.
MDAS, Modified Dental Anxiety Scale; SEM, structural equation model; MLR, robust maximum likelihood.

The MDAS supported configural, metric, and scalar invariance across smoking groups (Appendix Table 8). For the GAD-7, practical fit changes were small (ΔCFI ≤ 0.010), supporting sufficient comparability for use as an adjusted latent covariate, although the scaled chi-square difference tests did not support exact invariance (Appendix Table 9). Multi-group SEM did not indicate clear between-group differences in structural paths or indirect and total associations (Appendix Table 10). Estimates among current smokers were less precise.

## Discussion

In this cross-sectional analysis of the Northern Finland Birth Cohort 1966, the prespecified pattern of associations was observed across behavioural, periodontal, and systemic-risk domains. Higher DA was associated with more delayed/problem-oriented oral healthcare attendance and poorer home oral care, which were associated with greater periodontal disease burden. Greater periodontal disease burden was, in turn, associated with higher cardiovascular risk. The overall association between DA and cardiovascular risk was small, as was the decomposed indirect association through the measured behavioural and periodontal domains, while a remaining direct association was also observed.

The principal contribution of the present study is that associations previously examined largely within separate studies were evaluated together in a single prespecified empirical model centred on DA. Associations of DA with avoidance and poorer oral health-related behaviours are well established [2, 5, 6], as are associations between periodontal disease and cardiovascular disease [10]. Population-based studies have also linked DA with poorer clinical oral health, although findings for periodontal outcomes have not been entirely consistent [34, 35]. The previous conceptual mapping review showed that systemic health has rarely been integrated into DA-focused frameworks [4], and proposed links between dental fear, oral health, and chronic disease risk have rarely been evaluated within an integrated population-based model [36]. The present study extends this literature by operationalizing behavioural, periodontal, and systemic-risk domains within one model and testing the prespecified cross-domain associations.

The extension to the systemic-risk domain warrants cautious interpretation. Greater periodontal disease burden was associated with higher cardiovascular risk, and a small overall association was observed between DA and cardiovascular risk. Because the cardiovascular outcome represented estimated risk rather than incident disease [19, 20], these findings do not establish DA as an important cardiovascular risk factor or a causal oral–systemic sequence. Smoking-stratified analyses did not reveal a consistent pattern of between-group differences, although estimates among current smokers were less precise. The small decomposed indirect association and remaining direct association further suggest that the present operationalization captured only part of the broader association. The remaining association may reflect unmeasured or incompletely represented domains, including continuity of oral healthcare, cumulative treatment history, broader health behaviours, social or healthcare-access vulnerabilities, or shared upstream determinants. These possibilities remain hypothetical, and marker relevance should not be conflated with causal explanation [37]. From a conceptual-model perspective, the measured behavioural–periodontal route may therefore represent only one component of a broader structure requiring further refinement.

Taken together, these findings broaden the relevance of DA beyond chairside management. They provide a rationale for examining whether DA may have value as a measurable oral health-specific psychosocial marker of vulnerability within broader patterns of oral health-related behaviour and disease burden, rather than as a cardiovascular risk predictor. Because DA can be assessed using brief validated instruments such as the MDAS, this hypothesis is empirically testable in population-based research. Incorporating brief DA assessments into routine health examinations could provide opportunities to identify individuals who may benefit from additional support. Whether DA provides useful information for identifying such vulnerability or improving care pathways remains to be established prospectively.

Strengths include the large population-based cohort and the combination of self-reported psychosocial and behavioural measures with clinically assessed periodontal status and a validated Framingham-based cardiovascular risk score within the same survey wave, reducing reliance on a single measurement source. The conceptual framework preceded the analysis, and the operational model was specified on a theory-informed basis rather than derived from observed associations. Modelling dental and general anxiety as latent constructs also allowed measurement error to be represented explicitly.

Several limitations define the scope of interpretation. First, the cross-sectional design does not establish temporal ordering, and the decomposed indirect associations should be interpreted as structural decompositions of association rather than evidence of causal mediation. Although the hypothesized ordering was theory-informed, reverse or bidirectional relationships and alternative causal structures cannot be excluded. In particular, conditioning on intermediate variables may introduce collider stratification bias if these variables are influenced by unmeasured causes of subsequent outcomes. The present data cannot determine whether such structures are operating, and the estimated pathways should not be interpreted as uniquely identified causal pathways. Residual confounding by unmeasured behavioural, metabolic, social, or healthcare-related factors also remains possible. Second, the behavioural domains were represented by pragmatic theory-based proxies derived from available cohort variables rather than validated measures of DA-related attendance or self-care. The delayed/problem-oriented attendance index intentionally combined distinct aspects of attendance rather than functioning as a reflective multi-item scale, and these indicators may have captured only part of the broader behavioural processes represented in the conceptual model. Periodontal disease burden was based on the percentage of sites with probing pocket depth ≥4 mm and did not capture all dimensions of periodontitis, while cardiovascular risk was represented by a Framingham-based risk score rather than incident cardiovascular disease. Finally, participation-related selection bias is possible, and restriction to 46-year-old members of a single Northern Finnish birth cohort limits generalizability to other age groups, populations, and healthcare settings.

In conclusion, this study provides the first population-based empirical evaluation of the consequences component of a previously proposed conceptual model, operationalized across oral health-related behaviour, periodontal disease burden, and distal cardiovascular risk. The hypothesized associations were observed across these domains, providing preliminary support for this empirically testable representation of the model rather than confirmation of a causal pathway. Longitudinal studies with broader operationalization of behavioural and healthcare-use domains are needed to establish temporal ordering and to examine whether DA may have value as a measurable oral health-specific psychosocial marker of vulnerability.

## Author Contributions

Mika Kajita contributed to conception, design, data curation, formal analysis, funding acquisition, validation, visualization, interpretation, drafted the manuscript, and critically revised the manuscript. Akie Yada contributed to design, methodology, funding acquisition, validation, interpretation, and critically revised the manuscript. Pekka Ylöstalo contributed to data acquisition, resources, validation, interpretation, and critically revised the manuscript. Vesa Pohjola contributed to conception, validation, interpretation, and critically revised the manuscript. Kirsi Sipilä contributed to data acquisition, project administration, interpretation, and critically revised the manuscript. Gerald Humphris contributed to design, interpretation, and critically revised the manuscript. Juha Auvinen contributed to data acquisition, resources, interpretation, and critically revised the manuscript. Satu Lahti contributed to conception, supervision, validation, interpretation, and critically revised the manuscript. All authors gave their final approval and agreed to be accountable for all aspects of the work.

## Acknowledgments

The authors thank all cohort members, researchers and NFBC project center personnel who participated in the NFBC data collections.

Generative AI (ChatGPT, OpenAI) was used as described in the Methods section. All AI-assisted outputs were critically reviewed, verified, and revised as appropriate by the authors. The authors take full responsibility for the scientific content, analyses, interpretation, and final manuscript.

## Ethics Approval and Consent to Participate

The study was conducted in accordance with the Declaration of Helsinki. The 46-year follow-up of the Northern Finland Birth Cohort 1966 (NFBC1966) was approved by the Ethics Committee of the Northern Ostrobothnia Hospital District. Written informed consent was obtained from all participants.

## Data Availability Statement

The individual-level NFBC1966 data used in this study are not publicly available because their use is governed by participants’ written informed consent and applicable data protection requirements. Access may be requested for research purposes from the University of Oulu Infrastructure for Population Studies through its electronic material request process. Data use is subject to approval by the NFBC project center and compliance with the EU General Data Protection Regulation (EU 2016/679) and the Finnish Data Protection Act. Further information is available from the NFBC project center and the NFBC website (www.oulu.fi/nfbc).

The SPSS and Mplus syntax files used for data preparation and analysis, together with the variable dictionary, smoking-variable derivation file, and analysis-output crosswalk, are publicly available in Zenodo at (https://doi.org/10.5281/zenodo.21447346). The Zenodo repository does not contain individual-level NFBC1966 data or the restricted analysis dataset.

## Abbreviations

MDAS: Modified Dental Anxiety Scale
CFI: comparative fit index
TLI: Tucker–Lewis index
RMSEA: root mean square error of approximation
SRMR: standardized root mean square residual

## Reference

1. Silveira ER, Cademartori MG, Schuch HS, Armfield JA, Demarco FF. Estimated prevalence of dental fear in adults: A systematic review and meta-analysis. Journal of Dentistry. 2021;108:103632.

2. Armfield JM. What goes around comes around: revisiting the hypothesized vicious cycle of dental fear and avoidance. Community Dent Oral Epidemiol. 2013;41:279–87.

3. Seligman LD, Hovey JD, Chacon K, Ollendick TH. Dental anxiety: An understudied problem in youth. Clin Psychol Rev. 2017;55:25–40.

4. Kajita M, Pohjola V, Humphris G, Lahti S. Dental Anxiety as a Potential Bottleneck in Oral-Systemic Health Pathways: A Conceptual Mapping Review of Review Articles. Dent J (Basel). 2026;14.

5. Liinavuori A, Tolvanen M, Pohjola V, Lahti S. Longitudinal interrelationships between dental fear and dental attendance among adult Finns in 2000-2011. Community Dent Oral Epidemiol. 2019;47:309–15.

6. Crocombe LA, Broadbent JM, Thomson WM, Brennan DS, Slade GD, Poulton R. Dental visiting trajectory patterns and their antecedents. J Public Health Dent. 2011;71:23–31.

7. Watt RG, Sheiham A. Integrating the common risk factor approach into a social determinants framework. Community Dent Oral Epidemiol. 2012;40:289–96.

8. Peres MA, Macpherson LMD, Weyant RJ, Daly B, Venturelli R, Mathur MR, et al. Oral diseases: a global public health challenge. Lancet. 2019;394:249–60.

9. Botelho J, Mascarenhas P, Viana J, Proença L, Orlandi M, Leira Y, et al. An umbrella review of the evidence linking oral health and systemic noncommunicable diseases. Nat Commun. 2022;13:7614.

10. Herrera D, Sanz M, Shapira L, Brotons C, Chapple I, Frese T, et al. Association between periodontal diseases and cardiovascular diseases, diabetes and respiratory diseases: Consensus report of the Joint Workshop by the European Federation of Periodontology (EFP) and the European arm of the World Organization of Family Doctors (WONCA Europe). J Clin Periodontol. 2023;50:819–41.

11. Baker SR, Heaton LJ, McGrath C. Evolution and development of methodologies in social and behavioural science research in relation to oral health. Community Dentistry and Oral Epidemiology. 2023;51:46–57.

12. Celeste RK, Colvara BC, Rech RS, Reichenheim ME, Bastos JL. Challenges in operationalizing conceptual models in aetiological research. Community Dentistry and Oral Epidemiology. 2023;51:58–61.

13. University of Oulu. Northern Finland Birth Cohort 1966 (Version 1). University of Oulu; 2020.

14. Nordström T, Miettunen J, Auvinen J, Ala-Mursula L, Keinänen-Kiukaanniemi S, Veijola J, et al. Cohort Profile: 46 years of follow-up of the Northern Finland Birth Cohort 1966 (NFBC1966). International Journal of Epidemiology. 2021;50:1786–7j.

15. Humphris GM, Morrison T, Lindsay SJ. The Modified Dental Anxiety Scale: validation and United Kingdom norms. Community Dent Health. 1995;12:143–50.

16. Humphris GM, Freeman R, Campbell J, Tuutti H, D’Souza V. Further evidence for the reliability and validity of the Modified Dental Anxiety Scale. Int Dent J. 2000;50:367–70.

17. Armfield JM, Stewart JF, Spencer AJ. The vicious cycle of dental fear: exploring the interplay between oral health, service utilization and dental fear. BMC Oral Health. 2007;7:1.

18. Tegelberg P, Tervonen T, Knuuttila M, Jokelainen J, Keinänen-Kiukaanniemi S, Auvinen J, et al. Association of hyperglycaemia with periodontal status: Results of the Northern Finland Birth Cohort 1966 study. Journal of Clinical Periodontology. 2021;48:25–37.

19. Keto J, Ventola H, Jokelainen J, Linden K, Keinänen-Kiukaanniemi S, Timonen M, et al. Cardiovascular disease risk factors in relation to smoking behaviour and history: a population-based cohort study. Open Heart. 2016;3:e000358.

20. D’Agostino RB, Sr., Vasan RS, Pencina MJ, Wolf PA, Cobain M, Massaro JM, et al. General cardiovascular risk profile for use in primary care: the Framingham Heart Study. Circulation. 2008;117:743–53.

21. Spitzer RL, Kroenke K, Williams JB, Löwe B. A brief measure for assessing generalized anxiety disorder: the GAD-7. Arch Intern Med. 2006;166:1092–7.

22. Dadalti MT, Cunha AJ, Souza TG, Silva BA, Luiz RR, Risso PA. Anxiety about dental treatment - a gender issue. Acta Odontol Latinoam. 2021;34:195–200.

23. Stein Duker LI, Grager M, Giffin W, Hikita N, Polido JC. The Relationship between Dental Fear and Anxiety, General Anxiety/Fear, Sensory Over-Responsivity, and Oral Health Behaviors and Outcomes: A Conceptual Model. Int J Environ Res Public Health. 2022;19:2380.

24. Halonen H, Nissinen J, Lehtiniemi H, Salo T, Riipinen P, Miettunen J. The Association Between Dental Anxiety And Psychiatric Disorders And Symptoms: A Systematic Review. Clin Pract Epidemiol Ment Health. 2018;14:207–22.

25. Khaing W, Vallibhakara SA, Attia J, McEvoy M, Thakkinstian A. Effects of education and income on cardiovascular outcomes: A systematic review and meta-analysis. Eur J Prev Cardiol. 2017;24:1032–42.

26. Emdin CA, Odutayo A, Wong CX, Tran J, Hsiao AJ, Hunn BH. Meta-Analysis of Anxiety as a Risk Factor for Cardiovascular Disease. Am J Cardiol. 2016;118:511–9.

27. Kallio A, Suominen A, Tolvanen M, Rantavuori K, Jussila H, Karlsson L, et al. Concurrent changes in dental anxiety and smoking in parents of the FinnBrain Birth Cohort Study. Eur J Oral Sci. 2023;131:e12912.

28. Huxley RR, Woodward M. Cigarette smoking as a risk factor for coronary heart disease in women compared with men: a systematic review and meta-analysis of prospective cohort studies. Lancet. 2011;378:1297–305.

29. Little RJA. A Test of Missing Completely at Random for Multivariate Data with Missing Values. Journal of the American Statistical Association. 1988;83:1198–202.

30. Hu Lt, Bentler PM. Cutoff criteria for fit indexes in covariance structure analysis: Conventional criteria versus new alternatives. Structural Equation Modeling: A Multidisciplinary Journal. 1999;6:1–55.

31. Kline RB. Principles and practice of structural equation modeling, 4th ed. New York, NY, US: Guilford Press; 2016. xvii, 534-xvii, p.

32. Cheung GW, Rensvold RB. Evaluating Goodness-of-Fit Indexes for Testing Measurement Invariance. Structural Equation Modeling: A Multidisciplinary Journal. 2002;9:233–55.

33. Humphris GM, Newton JT. Is the Modified Dental Anxiety Scale (MDAS) a Single or Two Construct Measure? A Theoretical and Pragmatic Perspective. Dent J (Basel). 2025;13:68.

34. Ng SK, Leung WK. A community study on the relationship of dental anxiety with oral health status and oral health-related quality of life. Community Dent Oral Epidemiol. 2008;36:347–56.

35. Armfield JM, Slade GD, Spencer AJ. Dental fear and adult oral health in Australia. Community Dent Oral Epidemiol. 2009;37:220–30.

36. Beaudette JR, Fritz PC, Sullivan PJ, Ward WE. Oral Health, Nutritional Choices, and Dental Fear and Anxiety. Dent J (Basel). 2017;5.

37. Schooling CM, Jones HE. Clarifying questions about “risk factors”: predictors versus explanation. Emerg Themes Epidemiol. 2018;15:10.

