## Appendix for "Dental Anxiety in Oral–Systemic Health: Empirical Evaluation of a Published Conceptual Model"

**Appendix Table 1. Construction of composite variables used in the SEM**

| **Composite** | **Constituent variable** | **Original coding** | **Recoding used in the analysis** | **Final composite rule** | **Interpretation** |
| --- | --- | --- | --- | --- | --- |
| Delayed/problem-oriented attendance index | Time since last dental visit (K24) | 1 = <1 year; 2 = 1–2 years; 3 = 3–4 years; 4 = 5 years; 5 = >5 years; 6 = cannot remember | 1 → 0.00; 2 → 0.25; 3 → 0.50; 4 → 0.75; 5 → 1.00; 6 → missing |  | Higher score = longer time since last visit |
|  | Pain at the last dental visit (K23 → Lastvisit_pain) | K23: 1 = Yes; 2 = No | Reverse-coded to Lastvisit_pain (1 = No; 2 = Yes); 1/2 coding retained. |  | Higher score = pain at the last visit |
|  | Delayed/problem-oriented dental attendance index | — | — | Mean of available K24_scaled and Lastvisit_pain values | Higher score = greater attendance burden |
| Home care index | Floss use | 1 = never or hardly ever; 2 = every/almost every day; 3 = occasionally during the week | 1 → 0, 2/3 → 1 |  | Higher score = better home care |
|  | Interdental brush use | 1 = never or hardly ever; 2 = every/almost every day; 3 = occasionally during the week | 1 → 0, 2/3 → 1 |  | Higher score = better home care |
|  | Toothbrushing frequency | 1 = once/day; 2 = twice/day; 3 = more than twice/day | 1 → 0, 2/3 → 1 |  | Higher score = better home care |
|  | Skipping toothbrushing due to fatigue/lack of motivation | 1 = rarely or never; 2 = every/almost every day; 3 = occasionally during the week | reverse-coded: 1 → 1, 2/3 → 0 |  | Higher score = less skipping / better home care |
|  | Home care index | — | — | Mean of the four binary items, calculated when at least 3 of 4 items were available | Higher score = better home care |

**Appendix Table 2. Descriptive statistics for MDAS and GAD-7 item responses**

| **Construct** | **Item** | **N** | **Scale range** | **Mean** | **SD** | **Skewness** | **Kurtosis** |
| --- | --- | --- | --- | --- | --- | --- | --- |
| MDAS | MDAS1 | 1872 | 1–5 | 1.5 | 0.84 | 2.08 | 4.68 |
| MDAS | MDAS2 | 1872 | 1–5 | 1.7 | 0.89 | 1.6 | 2.81 |
| MDAS | MDAS3 | 1872 | 1–5 | 2.21 | 1.01 | 1.05 | 0.94 |
| MDAS | MDAS4 | 1872 | 1–5 | 1.73 | 0.89 | 1.35 | 1.85 |
| MDAS | MDAS5 | 1872 | 1–5 | 2.12 | 0.97 | 0.97 | 0.85 |
| GAD-7 | Feeling nervous, anxious, or on edge | 1692 | 1–4 | 1.44 | 0.64 | 1.6 | 3.25 |
| GAD-7 | Not being able to stop or control worrying | 1693 | 1–4 | 1.32 | 0.59 | 2.06 | 4.84 |
| GAD-7 | Worrying too much about different things | 1693 | 1–4 | 1.43 | 0.64 | 1.57 | 2.79 |
| GAD-7 | Trouble relaxing | 1692 | 1–4 | 1.42 | 0.64 | 1.66 | 3.11 |
| GAD-7 | Being so restless that it is hard to sit still | 1692 | 1–4 | 1.14 | 0.43 | 3.75 | 17.21 |
| GAD-7 | Becoming easily annoyed or irritable | 1691 | 1–4 | 1.49 | 0.62 | 1.17 | 1.71 |
| GAD-7 | Feeling afraid as if something awful might happen | 1695 | 1–4 | 1.17 | 0.45 | 3.1 | 11.82 |

**Abbreviations:** MDAS, Modified Dental Anxiety Scale; GAD-7, Generalized Anxiety Disorder 7-item scale; SD, standard deviation. MDAS items were scored from 1 to 5. GAD-7 items are shown using a 1–4 coding format corresponding to the original 0–3 response options. Item-specific differences in N reflect missing responses.

**Appendix Table 3. Distribution of components contributing to the composite indices**

| **Indicator** | **Category or scaled category** | **n (valid %)** | **Missing, n (%)** |
| --- | --- | --- | --- |
| Pain at the last dental visit |  |  | 51 (2.7) |
|  | No | 1280 (68.4) |  |
|  | Yes | 592 (31.6) |  |
| Time since last dental visit |  |  | 59 (3.1) |
|  | <1 year | 1074 (57.6) |  |
|  | 1–2 years | 520 (27.9) |  |
|  | 3–4 years | 181 (9.7) |  |
|  | 5 years | 27 (1.4) |  |
|  | >5 years | 62 (3.3) |  |
| Dental floss use |  |  | 87 (4.5) |
|  | Never or hardly ever | 777 (42.3) |  |
|  | Every/almost every day | 361 (19.7) |  |
|  | Occasionally during the week | 698 (38.0) |  |
| Interdental brush use |  |  | 142 (7.4) |
|  | Never or hardly ever | 1573 (88.3) |  |
|  | Every/almost every day | 90 (5.1) |  |
|  | Occasionally during the week | 118 (6.6) |  |
| Toothbrushing frequency |  |  | 91 (4.7) |
|  | Once/day | 543 (29.6) |  |
|  | Twice/day | 1234 (67.4) |  |
|  | More than twice/day | 55 (3.0) |  |
| Skipping toothbrushing due to fatigue/lack of motivation |  |  | 52 (2.7) |
|  | Rarely or never | 1513 (80.9) |  |
|  | Every/almost every day | 20 (1.1) |  |
|  | Occasionally during the week | 338 (18.1) |  |

Appendix Table 4**.** Spearman correlations among components of the delayed/problem-oriented dental attendance index

| **Variable** | **1** | **2** | **3** |
| --- | --- | --- | --- |
| 1. Time since the last dental visit | — | 0.096** | 0.610** |
| 2. Pain at the last dental visit | 0.096** | — | 0.828** |
| 3. Delayed/problem-oriented dental attendance index | 0.610** | 0.828** | — |

**Notes.** Values are Spearman’s rho. The delayed/problem-oriented dental attendance index combined time since the last dental visit and pain at the last dental visit. Higher values indicate greater delayed/problem-oriented dental attendance. Pairwise sample sizes ranged from 1,864 to 1,872.
****p < 0.001.**

Appendix Table 5. Missingness in variables included in the main structural equation model

| **Variable** | **Observed n** | **Missing n (%)** | **Observed mean** | **Observed SD** |
| --- | --- | --- | --- | --- |
| MDAS1 | 1872 | 51 (2.7) | 1.502 | 0.837 |
| MDAS2 | 1872 | 51 (2.7) | 1.696 | 0.894 |
| MDAS3 | 1872 | 51 (2.7) | 2.210 | 1.012 |
| MDAS4 | 1872 | 51 (2.7) | 1.728 | 0.890 |
| MDAS5 | 1872 | 51 (2.7) | 2.120 | 0.965 |
| GAD1 | 1692 | 231 (12.0) | 1.44 | 0.638 |
| GAD2 | 1693 | 230 (12.0) | 1.32 | 0.594 |
| GAD3 | 1693 | 230 (12.0) | 1.43 | 0.640 |
| GAD4 | 1692 | 231 (12.0) | 1.42 | 0.642 |
| GAD5 | 1692 | 231 (12.0) | 1.14 | 0.429 |
| GAD6 | 1691 | 232 (12.1) | 1.49 | 0.618 |
| GAD7 | 1695 | 228 (11.9) | 1.17 | 0.450 |
| Periodontal disease burden, % sites with PPD ≥4 mm | 1897 | 26 (1.4) | 2.755 | 6.264 |
| Framingham cardiovascular risk score | 1744 | 179 (9.3) | 6.372 | 4.787 |
| Delayed/problem-oriented dental attendance index | 1872 | 51 (2.7) | 0.742 | 0.274 |
| Home care index | 1791 | 132 (6.9) | 0.557 | 0.256 |
| Sex | 1923 | 0 (0.0) | — | — |
| Education | 1859 | 64 (3.3) | — | — |
| Smoking status | 1861 | 62 (3.2) | — | — |

**Note.** The analytic dataset included 1,923 participants. MDAS, Modified Dental Anxiety Scale; GAD-7, Generalized Anxiety Disorder 7-item scale; PPD, probing pocket depth; SD, standard deviation. Means and standard deviations are shown for continuous or ordinal variables used as continuous indicators in the SEM; they are not shown for categorical variables. Little’s MCAR test was statistically significant, χ²(298) = 441.65, p < .001, indicating that the strict MCAR assumption was not supported. However, EM-estimated means and standard deviations were highly similar to the observed estimates. Missing data in the structural equation model were handled using full information maximum likelihood.

Appendix Table 6. Comparison of one-factor CFA models for the Modified Dental Anxiety Scale (MDAS)

Panel A. Model fit indices

| **Model** | **N** | **χ² (df)** | **CFI / TLI** | **RMSEA (90% CI)** | **SRMR** |
| --- | --- | --- | --- | --- | --- |
| One-factor model | 1872 | 235.58 (5) | 0.951 / 0.902 | 0.157 (0.140–0.174) | 0.033 |
| One-factor model with correlated residuals (MDAS1 WITH MDAS2) | 1872 | 35.52 (4) | 0.993 / 0.983 | 0.065 (0.046–0.085) | 0.013 |

Panel B. Standardized factor loadings (STDYX)

| **Item** | **One-factor model** | **One-factor model with correlated residuals** |
| --- | --- | --- |
| MDAS1 | 0.898 | 0.813 |
| MDAS2 | 0.926 | 0.852 |
| MDAS3 | 0.848 | 0.904 |
| MDAS4 | 0.755 | 0.775 |
| MDAS5 | 0.714 | 0.765 |

Residual correlation added in the revised model

| **Parameter** | **Estimate (STDYX)** | **p-value** |
| --- | --- | --- |
| MDAS1 WITH MDAS2 | 0.544 | <0.001 |

**Abbreviations:** CFA, confirmatory factor analysis; MDAS, Modified Dental Anxiety Scale; CFI, Comparative Fit Index; TLI, Tucker–Lewis Index; RMSEA, Root Mean Square Error of Approximation; SRMR, Standardized Root Mean Square Residual.

**Note:** Both models were estimated using robust maximum likelihood (MLR). The correlated residual between MDAS1 and MDAS2 was added based on the modification indices from the initial one-factor model.

Appendix Table 7. One-factor CFA model for the Generalized Anxiety Disorder scale (GAD-7)

**Panel A. Model fit indices**

| **Model** | **N** | **χ² (df)** | **CFI / TLI** | **RMSEA (90% CI)** | **SRMR** |
| --- | --- | --- | --- | --- | --- |
| One-factor model | 1699 | 88.00 (14) | 0.969 / 0.953 | 0.056 (0.045–0.067) | 0.029 |

**Panel B. Standardized factor loadings (STDYX)**

| **Item** | **Standardized loading** |
| --- | --- |
| GAD1 | 0.779 |
| GAD2 | 0.781 |
| GAD3 | 0.810 |
| GAD4 | 0.782 |
| GAD5 | 0.600 |
| GAD6 | 0.686 |
| GAD7 | 0.626 |

**Abbreviations:** CFA, confirmatory factor analysis; GAD-7, Generalized Anxiety Disorder 7-item scale; CFI, Comparative Fit Index; TLI, Tucker–Lewis Index; RMSEA, Root Mean Square Error of Approximation; SRMR, Standardized Root Mean Square Residual.
**Note:** The model was estimated using robust maximum likelihood (MLR).

Appendix Table 8. Measurement invariance of the Modified Dental Anxiety Scale across smoking groups

| **Model / constraint** | **n** | **χ²(df)** | **CFI / TLI** | **RMSEA (90% CI) / SRMR** | **Difference test and interpretation** |
| --- | --- | --- | --- | --- | --- |
| Single-group CFA: non-current smokers | 1401 | 32.842 (4) | 0.993 / 0.982 | 0.072 (0.050–0.095) / 0.015 | Good fit |
| Single-group CFA: current smokers | 410 | 8.549 (4) | 0.995 / 0.988 | 0.053 (0.000–0.102) / 0.012 | Good fit |
| Configural invariance | 1811 | 40.273 (8) | 0.993 / 0.982 | 0.067 (0.047–0.088) / 0.015 | Acceptable configural model |
| Metric invariance | 1811 | 42.848 (12) | 0.993 / 0.988 | 0.053 (0.037–0.071) / 0.018 | vs configural: Δχ² = 2.364, Δdf = 4, p = 0.669; metric invariance supported |
| Scalar invariance | 1811 | 51.574 (16) | 0.992 / 0.990 | 0.050 (0.035–0.065) / 0.018 | vs metric: Δχ² = 7.046, Δdf = 4, p = 0.134; scalar invariance supported. vs configural: Δχ² = 8.652, Δdf = 8, p = 0.373 |

**Abbreviations:** CFI, comparative fit index; TLI, Tucker–Lewis index; RMSEA, root mean square error of approximation; SRMR, standardized root mean square residual. Smoking groups were defined from the original smoking-status variable in the source dataset (coded 1–5) and dichotomized as non-current smokers (1–3) and current smokers (4–5). A correlated residual between MDAS1 and MDAS2 was specified in all models. Invariance was evaluated sequentially at the configural, metric, and scalar levels using MLR estimation. Chi-square difference tests are the Mplus scaled difference tests for MLR.

Appendix Table 9. Single-group and multi-group CFA results for the GAD-7 across smoking groups

| **Model / group or constraint** | **n** | **χ²(df)** | **CFI / TLI** | **RMSEA (90% CI) / SRMR** | **Interpretation** |
| --- | --- | --- | --- | --- | --- |
| Single-group CFA: non-current smokers | 1312 | 75.061 (14) | 0.964 / 0.946 | 0.058 (0.045–0.071) / 0.030 | Acceptable fit |
| Single-group CFA: current smokers | 358 | 25.467 (14) | 0.984 / 0.977 | 0.048 (0.015–0.077) / 0.026 | Good fit |
| Configural invariance | 1670 | 106.761 (28) | 0.968 / 0.952 | 0.058 (0.047–0.070) / 0.029 | Acceptable configural model |
| Metric invariance | 1670 | 126.547 (34) | 0.963 / 0.954 | 0.057 (0.047–0.068) / 0.058 | Compared with the configural model: Δχ² = 20.415, Δdf = 6, p = 0.002; chi-square difference test was significant, although changes in practical fit indices were modest (ΔCFI = -0.005, ΔRMSEA = -0.001, ΔSRMR = +0.029). |
| Scalar invariance | 1670 | 149.832 (40) | 0.956 / 0.953 | 0.057 (0.048–0.067) / 0.053 | Compared with the metric model: Δχ² = 24.348, Δdf = 6, p < 0.001; chi-square difference test was significant, although changes in practical fit indices were modest (ΔCFI = -0.007, ΔRMSEA = 0.000, ΔSRMR = -0.005). Compared with the configural model: Δχ² = 42.844, Δdf = 12, p < 0.001. |

**Abbreviations:** GAD-7, Generalized Anxiety Disorder-7; CFA, confirmatory factor analysis; CFI, comparative fit index; TLI, Tucker–Lewis index; RMSEA, root mean square error of approximation; SRMR, standardized root mean square residual. Smoking groups were defined as non-current smokers and current smokers. Multi-group CFA was estimated using MLR. Invariance decisions were based primarily on changes in practical fit indices.

Appendix Table 10. Multi-group structural equation model by smoking status: indirect, direct, and total associations of dental anxiety with cardiovascular risk score

| **Path / Effect** | **Non-current smokers Estimate (SE)** | **p** | **Current smokers Estimate (SE)** | **p** | **Group difference Estimate (SE)** | **p** |
| --- | --- | --- | --- | --- | --- | --- |
| **Structural paths (unstandardized)** |  |  |  |  |  |  |
| DA → Delayed/problem-oriented dental attendance | 0.078 (0.013) | <.001 | 0.061 (0.020) | 0.003 | -0.017 (0.024) | 0.474 |
| DA → Home care | -0.043 (0.011) | <.001 | -0.041 (0.019) | 0.029 | 0.002 (0.021) | 0.919 |
| DA → Periodontal disease burden | 0.574 (0.264) | 0.029 | 0.599 (0.641) | 0.349 | 0.025 (0.684) | 0.971 |
| Delayed/problem-oriented dental attendance → Periodontal disease burden | 0.853 (0.529) | 0.107 | 3.032 (1.600) | 0.058 | 2.180 (1.686) | 0.196 |
| Home care → Periodontal disease burden | -1.291 (0.620) | 0.037 | -3.379 (1.657) | 0.041 | -2.087 (1.730) | 0.228 |
| Periodontal disease burden → Cardiovascular risk | 0.026 (0.015) | 0.077 | 0.074 (0.038) | 0.049 | 0.048 (0.040) | 0.234 |
| DA → Cardiovascular risk (direct effect) | 0.306 (0.151) | 0.043 | -0.352 (0.338) | 0.297 | -0.658 (0.361) | 0.069 |
| **Indirect and total effects of DA on cardiovascular risk** | |  |  |  |  |  |
| Via periodontal disease burden only | 0.015 (0.011) | 0.161 | 0.044 (0.053) | 0.400 | 0.029 (0.053) | 0.581 |
| Via Delayed/problem-oriented dental attendance → periodontal disease burden | 0.002 (0.001) | 0.203 | 0.014 (0.010) | 0.172 | 0.012 (0.010) | 0.236 |
| Via home care → periodontal disease burden | 0.001 (0.001) | 0.184 | 0.010 (0.009) | 0.250 | 0.009 (0.009) | 0.326 |
| Total indirect effect | 0.018 (0.012) | 0.127 | 0.068 (0.059) | 0.250 | 0.050 (0.060) | 0.404 |
| Total effect | 0.324 (0.152) | 0.033 | -0.284 (0.340) | 0.404 | -0.608 (0.364) | 0.096 |

Group difference was defined as current smokers minus non-current smokers.

Note. Estimates are unstandardized coefficients from the smoking-stratified multi-group structural equation model estimated using robust maximum likelihood (MLR). Factor loadings for the MDAS and GAD-7 were constrained to equality across smoking groups, whereas item intercepts were freely estimated across groups. Latent means were fixed at zero in both groups. The focal structural paths involving dental anxiety were freely estimated in each group, while regression paths for generalized anxiety, sex, and education were constrained to equality across groups. The covariance between dental anxiety and generalized anxiety was also constrained to equality across groups. A residual covariance between MDAS1 and MDAS2 was specified in both groups. Group differences were calculated as current smokers minus non-current smokers. DA, dental anxiety; SE, standard error.
